# A validated hypoxia-inducible factor (HIF) signature across tissue compartments predicts clinical outcome in human lung fibrosis

**DOI:** 10.1101/2022.01.11.22269085

**Authors:** Yilu Zhou, Rob M. Ewing, Donna E. Davies, Yihua Wang, Mark G. Jones

**Author notes:** Correspondence should be addressed to M.G.J. (e- mail) or Y.W. Senior authors contributed equally to this work (Yihua Wang, Mark G. Jones).

## Abstract

We previously reported that oxidative stress drives pseudohypoxic hypoxia-inducible factor (HIF) pathway activation to promote pathogenetic collagen structure-function in human lung fibrosis (Brereton et al., 2022). Here, through bioinformatic studies we investigate HIF pathway activation status in patients with idiopathic pulmonary fibrosis (IPF) and whether this has prognostic significance. Applying a well-established HIF gene expression signature, we classified publicly available datasets into HIF score-high and score-low groups across multiple tissue compartments. The HIF scores in lung tissue, bronchoalveolar lavage (BAL) and peripheral blood mononuclear cells (PBMC) were increased in IPF patients and significantly correlated with an oxidative stress signature consistent with pseudohypoxic HIF pathway activation. A high HIF score in BAL and in PBMC was a strong independent predictor of mortality in multivariate analysis. Thus, a validated HIF gene signature predicts survival across tissue compartments in IPF and merits prospective study as a non-invasive biomarker of lung fibrosis progression.

## Introduction

We previously reported that altered collagen nano-architecture is a defining feature of idiopathic pulmonary fibrosis (IPF) that dysregulates extracellular matrix (ECM) structure-function to promote progressive lung fibrosis (1). We have recently extended these observations, identifying that hypoxia-inducible factor (HIF) pathway activation is required to promote pathologic pyridinlone collagen crosslinking and tissue stiffness by disproportionate induction of collagen-modifying enzymes relative to TGFβ-induced collagen fibril synthesis (2). Furthermore, we identified that this may occur via oxygen-independent (pseudohypoxic) mechanisms, including a decrease in factor inhibiting HIF (FIH) activity due to oxidative stress (2). Oxidative stress was increased in subpopulations of IPF fibroblasts whilst FIH activity was significantly reduced in fibroblasts from patients with lung fibrosis resulting in HIF activation under normoxic conditions. To further assess for HIF activity we applied a validated HIF/hypoxia metagene signature, identifying that HIF activity was increased at sites of active fibrogenesis in IPF tissue as well as within IPF mesenchymal cell populations. Intriguingly, in a small lung mesenchymal stromal RNAseq data set increased HIF pathway activation assessed via gene set variation analysis (GSVA) of this HIF signature was associated with disease progression. Here we sought to further investigate the potential utility of this validated HIF signature as a predictor of disease progression.

## Results

### A validated HIF score is increased across tissue compartments in patients with IPF

We applied a 15-gene expression signature (*ACOT7, ADM, ALDOA, CDKN3, ENO1, LDHA, MIF, MRPS17, NDRG1, P4HA1, PGAM1, SLC2A1, TPI1, TUBB6* and *VEGFA*), which has been validated across multiple populations (3-6) and which enables classification of HIF activity by calculating a HIF score for each sample using GSVA, to publicly available datasets (7). We firstly confirmed that the HIF score was significantly increased in the transcriptome of bulk lung tissues from patients with IPF compared to control tissues (Fig. 1A), and that this correlated (*R* = 0.89, *P* = 7.9×10^−15^) with an upregulated oxidative stress gene expression signature (Fig. 1B), findings consistent with our previous observation of increased pseudohypoxic HIF activity in IPF tissue (2).

**Figure 1.**
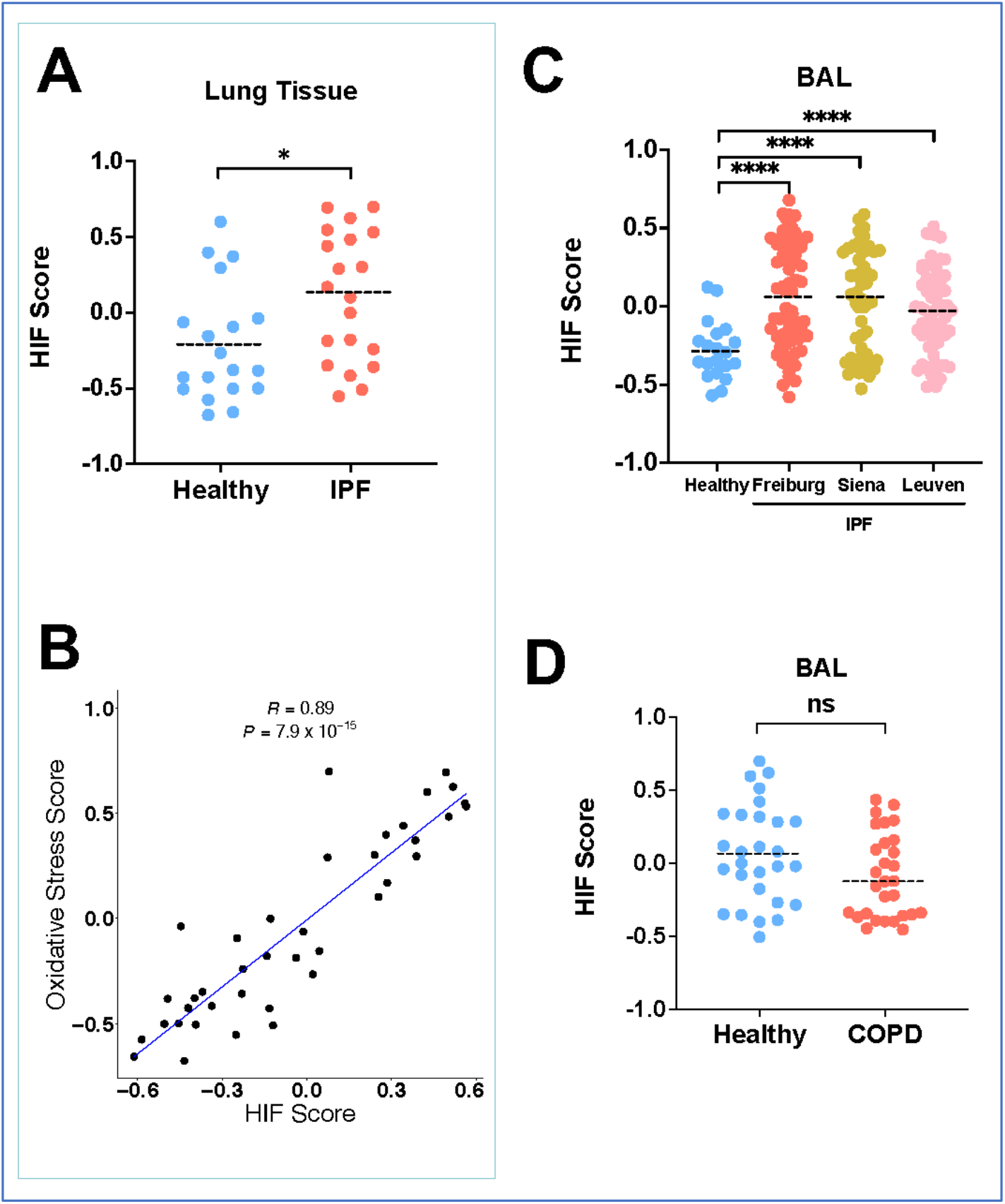
A validated HIF score is increased across tissue compartments in patients with IPF. (*A*) HIF GSVA scores from lung tissue from healthy controls and IPF patients (GSE92592). Data are mean. \**P* < 0.05 by unpaired *t*-test. (*B*) The scatter plot for the correlation between HIF score and oxidative stress score in IPF lung tissues (Pearson’s correlation *R* = 0.89 and *P* = 7.9×10^−15^). (*C*) HIF GSVA scores from bronchoalveolar lavage cells from 3 independent IPF cohorts (Freiburg, Siena and Leuven) compared to healthy donors (GSE70867). Data are mean. \*\*\*\**P* < 0.0001 by Dunnett’s multiple comparisons test. (*D*) HIF GSVA scores in BAL samples from patients with COPD compared to healthy smokers. Data are mean. ns (not significant) by unpaired *t*-test, *P* > 0.05.

We then investigated the HIF score calculated from the bronchoalveolar lavage (BAL) cell transcriptome of 176 patients from 3 independent IPF cohorts: Freiburg (Germany), Siena (Italy) and Leuven (Belgium) (8). Compared to healthy volunteers, the HIF score was significantly increased in patients with IPF across all 3 cohorts (Fig. 1C; *P* < 0.001), whilst there was no significant difference in the HIF score between smoker control and chronic obstructive pulmonary disease (COPD) patients (Fig. 1D). The HIF score in BAL samples also correlated with an oxidative stress gene expression signature (*R* = 0.48, *P* = 1.3×10^−11^; Figure 1-figure supplement 1). Together these findings identify that a validated HIF score is increased in both lung tissue and bronchoalveolar lavage cells from patients with IPF, and that this is associated with increased oxidative stress.

### The HIF score in BAL predicts mortality in patients with IPF

We next investigated whether the HIF score itself had prognostic value in this BAL cohort. Classifying patients into score-high and score-low groups based on unsupervised hierarchical clustering (Figure 2-figure supplement 1A; Fig. 2A), we identified that the HIF score significantly predicted survival in the whole IPF cohort (Fig. 2B; hazard ratio, HR: 5.4; *P* = 2.55×10^−7^), with comparable trends in each individual cohort (Figure 2-figure supplement 1B-D). A high HIF score was a strong independent predictor (HR: 4.1, *P* < 0.001) of mortality including in multivariate analysis with the physiological Gender, Age and Physiology (GAP) score that uses commonly measured clinical and physiologic variables to predict mortality in IPF (9) (Fig. 2C). Notably, the HIF score alone performed similarly to the GAP index (Fig. 2D), and when the HIF score and the GAP index were combined, the outcome prediction error rate was reduced, indicating a significant added value (Fig. 2D).

**Figure 2.**
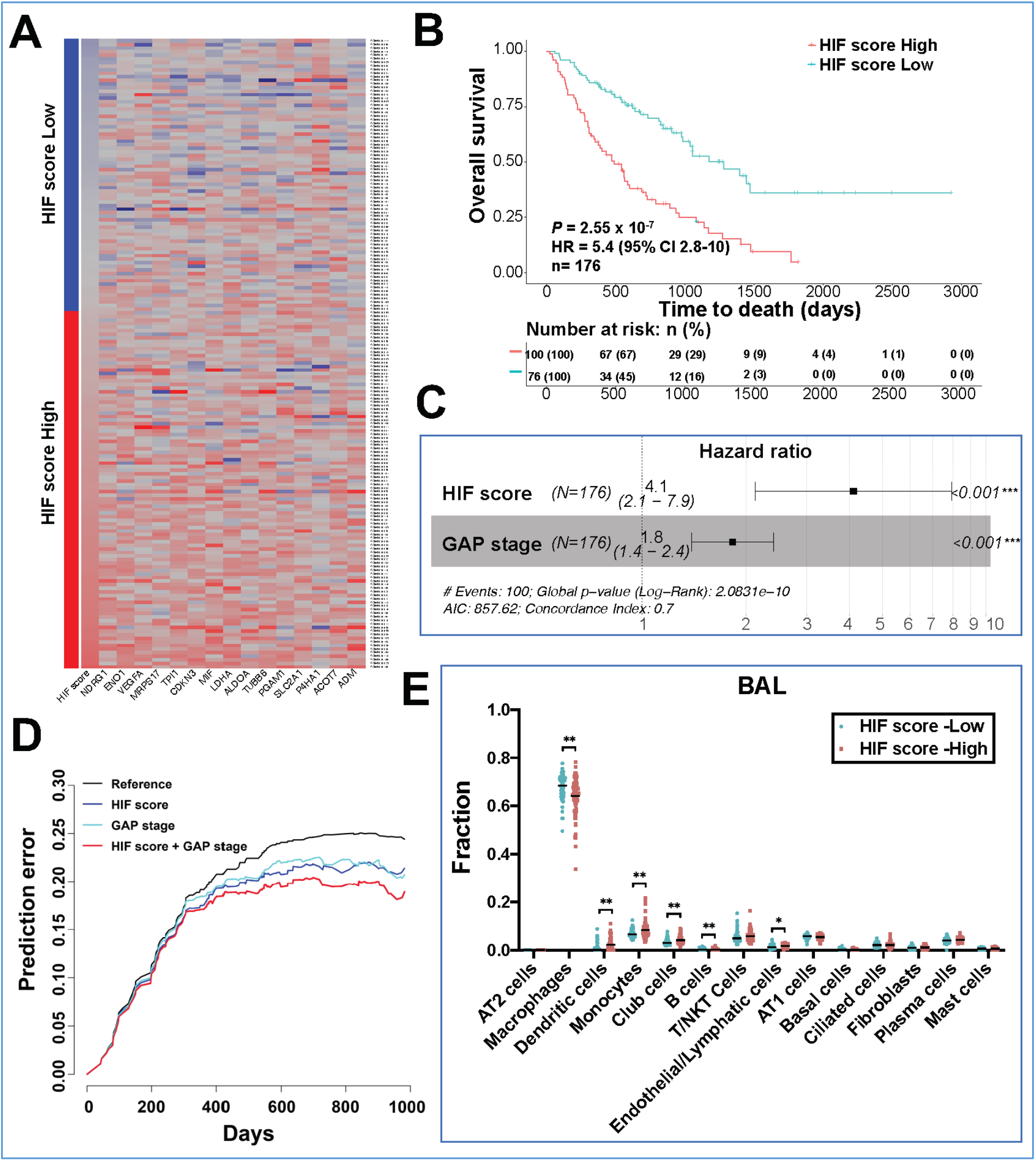
The HIF score in BAL predicts mortality in patients with IPF. (*A*) Heat map of the HIF score (*ACOT7, ADM, ALDOA, CDKN3, ENO1, LDHA, MIF, MRPS17, NDRG1, P4HA1, PGAM1, SLC2A1, TPI1, TUBB6* and *VEGFA*) from bronchoalveolar lavage cells of patients with IPF (GSE70867). Red indicates up-regulation and blue down-regulation. (*B*) Kaplan-Meier plot showing the overall survival in IPF patients with low *vs*. high HIF scores in BAL. Numbers below are *n* (%). *P* values, hazard ratio (HR), 95% confidence interval (CI) and patient number (*n*) are indicated. (*C*) Multivariate analysis in IPF patients. HR (hazard ratio), 95% CI (confidence interval), patient number (*n*) and *P* values are shown. (*D*) Prediction error curves (indicating mean squared error in predicting survival status) were calculated for the HIF score (blue) and Gender Age Physiology (GAP) index (green). Combining the HIF score with GAP score (red) reduced prediction errors and resulted in better prediction. (*E*) Cell deconvolution based on a signature matrix derived from the single-cell RNAseq found in Reyfman *et al* was used to determine cell compositions in BAL samples from IPF patients with low vs. high HIF score (GSE70867). \**P* < 0.05 by unpaired *t*-test. \*\**P* < 0.01.

To consider whether the change in HIF score might represent perturbations in cell types we used annotated lung scRNA-seq data (10) as a reference to perform cell type deconvolution analysis (CIBERSORTx) (11) of the bulk BAL cell gene profile. This identified macrophages (∼70%) as the major cell type in BAL (Figure 2-figure supplement 1E), a proportion consistent with the reported BAL macrophage differential count of this patient group (8). Comparing IPF patients with high HIF and low HIF scores identified a significant reduction in the macrophage proportion in HIF score high patients whilst the proportion of monocytes, B cells, dendritic cells, endothelial cells and club cells were significantly increased (Fig. 2E).

### The HIF score is increased in peripheral blood mononuclear cells (PBMC) from IPF patients

Although BAL provides direct insight into molecular events in the lung milieu, whole blood and peripheral blood mononuclear cells (PBMC) have the advantage of being a less invasive sampling method (12) and so we next applied the HIF score to PBMC transcriptome datasets. We first applied the HIF gene expression signature (3) to a PBMC microarray dataset (GSE38958) (13, 14) encompassing 45 healthy controls and 70 IPF patients, identifying that the HIF score was increased in PBMC samples from IPF patients compared to health controls (Fig. 3A; *P* < 0.05).

**Figure 3.**
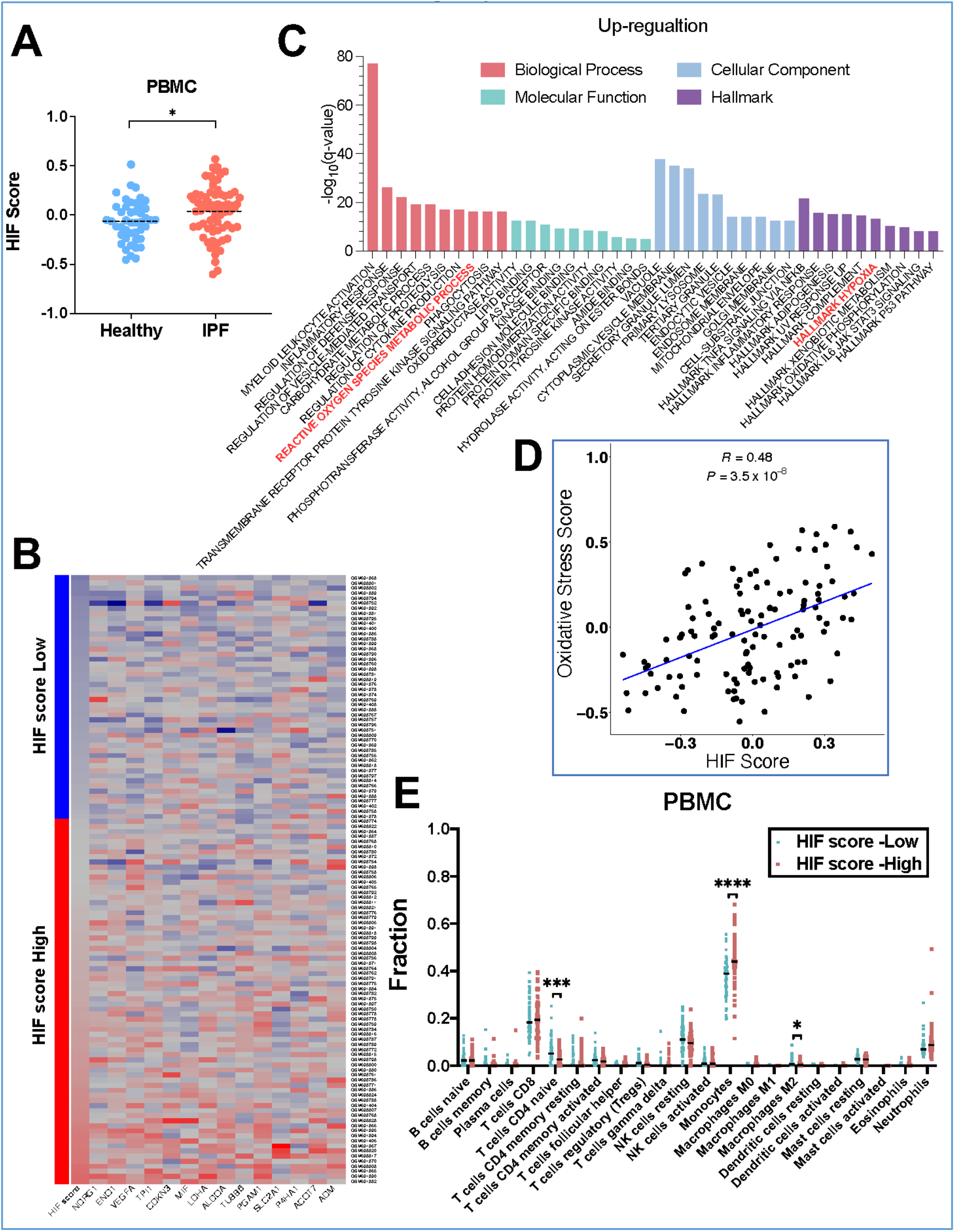
The HIF score is increased in PBMC from IPF patients. (*A*) HIF GSVA scores in PBMC samples from healthy controls and IPF patients (GSE38958). Data are mean. \**P* < 0.05 by unpaired *t*-test. (*B*) Heat map of the HIF score (*ACOT7, ADM, ALDOA, CDKN3, ENO1, LDHA, MIF, MRPS17, NDRG1, P4HA1, PGAM1, SLC2A1, TPI1, TUBB6* and *VEGFA*) in the microarray datasets GSE28042 and GSE27957. Red indicates up-regulation and blue down-regulation. (*C*) GO and pathway enrichment in HIF score high PBMC samples visualised on a bar chart clustered by molecular functions, cellular component, biological process and hallmark pathway. (*D*) The scatter plot for the correlation between HIF score and oxidative stress score in PBMC samples (Pearson’s correlation *R* = 0.48 and *P* = 3.5×10^−8^). (*E*) Cell deconvolution based on a signature matrix from Newman *et al* was used to determine cell compositions in PBMC samples from IPF patients with low vs. high HIF score (GSE38958). \**P* < 0.05. \*\*\**P* < 0.001. \*\*\*\**P* < 0.0001 by unpaired *t*-test.

We next calculated the HIF score in 2 PBMC microarray datasets of IPF patients with available longitudinal outcome data (GSE28042, Pittsburgh, n=75; and GSE27957, Chicago, n=45) (15, 16). Classifying patients into score-high and score-low groups (Figure 3-figure supplement 1A; Fig. 3B), we performed gene ontology (GO) and pathway enrichment analysis of differentially expressed genes (DEGs), identifying enrichment for “reactive oxygen species metabolic process” and “hallmark hypoxia” within the HIF score high group (Fig. 3C). Furthermore, applying the HIF signature or oxidative stress gene expression signatures to this dataset we identified a significant correlation, consistent with an increase in pseudohypoxic HIF activity in PBMCs (*R* = 0.48, *P* = 3.5×10^−8^; Fig. 3D).

To characterize the cell type composition within the PBMC RNA-seq data we next applied CIBERSORTx deconvolution using a leukocyte gene signature matrix developed by Newman and colleagues (17) as a reference (Figure 3-figure supplement 1B). Consistent with our observations in BAL samples, the proportion of monocytes identified in PBMCs was significantly increased in IPF patients with higher HIF scores (Fig. 3E; 44.4% *vs*. 37.1%; *P* < 0.0001) whilst the naïve CD4 T cell population and M2 Macrophages were decreased.

### The HIF score in PBMC from IPF patients predicts mortality

Finally, we investigated whether the HIF score had prognostic value in PBMC. We identified that a high HIF score predicted mortality in IPF patients from the Pittsburgh cohort (Figure 4-figure supplement 1A; HR: 13; *P* = 0.021), Chicago cohort (Figure 4-figure supplement 1B; HR: 4.3; *P* = 0.047) and the combined cohort (Chicago+ Pittsburgh) (Fig. 4A; HR: 7.2; *P* = 0.0073). A high HIF score in PBMC was also a strong independent predictor (HR: 5.9, *P* = 0.003) of mortality in multivariate analysis with gender and age (Fig. 4B). Thus, HIF activity assessed by a validated HIF metagene signature is a strong independent predictor of mortality across tissue compartments in patients with IPF.

**Figure 4.**
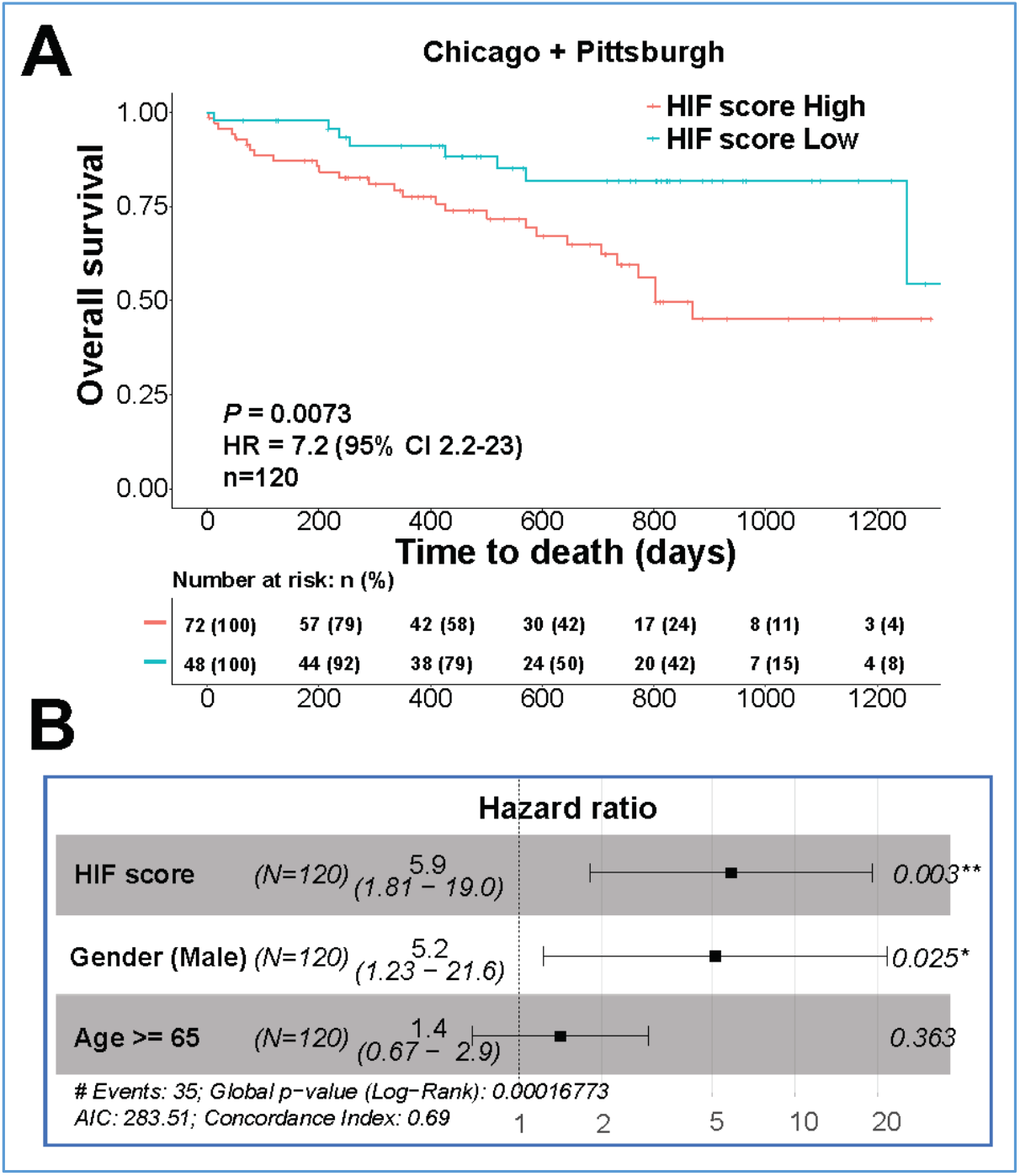
The HIF score in PBMC from IPF patients predicts mortality. (*A*) Kaplan-Meier plots showing the overall survival in IPF patients with low *vs*. high HIF scores in PBMC from the combined cohort (Chicago+ Pittsburg; GSE28042 and GSE27957). Numbers below are *n* (%). *P* values, hazard ratio (HR), 95% confidence interval (CI) and patient number (*n*) are indicated. (*B*) Multivariate analysis in IPF patients. HR (hazard ratio), 95% CI (confidence interval), patient number (*n*) and *P* values are shown.

## Discussion

We previously reported that altered collagen fibril nanoarchitecture is a core determinant of dysregulated ECM structure-function in human lung fibrosis (1), and that this dysregulation is promoted by pseudohypoxic HIF pathway activation (2). Here, we extend these observations to identify that HIF activity, as determined by a HIF gene signature, is increased in lung tissue, BAL, and PBMC samples from patients with IPF, that this strongly correlates with an oxidative stress signature consistent with pseudohypoxic HIF pathway activation, and that this is a strong independent predictor of mortality.

We selected a well-validated 15-gene expression HIF signature (3-6) to investigate HIF activity. In multiple studies this 15-gene signature has been identified to strongly correlate with independent hypoxia/HIF signatures (4-6). Furthermore, it was shown to be the best performer in a comprehensive study assessing the robustness of different hypoxia/HIF signatures (18). Importantly, it performed consistently across multiple pre-processing analytical pipelines (18), so supporting potential utility as a biomarker of disease activity. Our identification of consistent findings across tissue compartments in multiple patient cohorts is supportive of the need for prospective clinical studies to investigate the clinical utility of the HIF signature as a biomarker of disease activity. Although our study has focused upon patients with IPF, plausibly the 15-gene signature might have utility as a predictor of disease progression for any form of lung fibrosis.

Whilst hypoxia causes activation of the HIF pathway by inhibition of HIF prolyl hydroxylases that hydroxylate HIF-α and target it for degradation via the ubiquitin proteasome system, our recent study in IPF identified evidence of increased HIF activity under normoxic conditions (pseudohypoxic HIF activation) as a consequence of oxidative stress. Although we cannot exclude the possibility that hypoxia status contributed to HIF score in the cohorts studied, the HIF score was a strong independent predictor of mortality in multivariate analysiswith the physiological Gender, Age and Physiology (GAP) score, concordant with HIF pathway activation status being an independent biomarker of disease activity rather than simply a reflection of hypoxia status determined by disease severity. Additionally, we identified that the HIF score correlated with an oxidative stress gene expression signature across multiple tissue compartments, consistent with systemic oxidative stress modulating HIF pathway activity status (19).

Oxidative stress has been strongly implicated as an important profibrotic mechanism in the lungs, with N-acetylcysteine (NAC), a precursor of the anti-oxidant glutathione, proposed as a potential treatment for IPF. However, the role of NAC for IPF treatment remains controversial, with the PANTHER study identifying no benefit compared to placebo (20). Whilst NAC is not recommended as a treatment for IPF, meta-analyses continue to propose potential clinical utility (21). Further consideration is warranted into whether patients with a high HIF-score might benefit from NAC treatment. In support of the potential for stratified NAC treatment guided by HIF-score, post-hoc exploratory analysis of patients from the PANTHER study has proposed that in individuals with a polymorphism of the *TOLLIP* gene (TT rs3750920), NAC may be efficacious by decreasing TLR-4 oxidant dependent signaling (22).

We identified an increased monocyte proportions in BAL and PBMC samples with a high HIF score. A number of recent reports have proposed peripheral blood monocyte count as a predictor of IPF disease behaviour (23-25), with elevated monocyte count associated with increased risks of IPF progression, hospitalization, and mortality. Further investigation is required to confirm whether an increased HIF score reflects a dysregulated monocyte population and/or increased monocyte population. HIF activity is reported to regulate the development and function of hematopoietic progenitor/stem cells, with HIF pathway activation promoting metabolic and phenotypic reprogramming of myeloid cells (26). Consistent with these reports using a clinically relevant chronic intermittent hypoxia (CIH) model, Alvarez-Martins and colleagues were able to show an increase in monocytes upon CIH exposure (27), potentially via modulating monocyte differentiation, proliferation, and survival (28, 29). Furthermore, it has been recently reported (30) that monocytes from IPF patients are phenotypically distinct compared to age matched controls and this was associated with an increase in the level of macrophage colony-stimulating factor 1 (CSF-1) in serum. Notably, CSF-1 gene transcription has previously been reported (31) to be dependent upon HIF transcriptional activity.

In summary, we identify that a validated HIF score is increased across tissue compartments in patients with IPF, that this strongly correlates with oxidative stress consistent with pseudohypoxic HIF activation, and that an increase in this HIF score is a strong independent predictor of mortality. Prospective validation of these findings is warranted which could inform stratified approaches for the treatment of lung fibrosis.

## Methods

### Data preparation, analysis for Gene Ontology (GO) and pathway enrichment

Microarray data of BAL and PBMC samples are publicly available in NCBI in the Gene Expression Omnibus (GEO) with the accessions GSE70867 (8), GSE73395 (8), GSE38958 (13, 14), GSE28042 and GSE27957 (15, 16). Raw microarray files were downloaded and imported into Rstudio using the GEOquery (v2.56.0). The normalization of raw data depended on the analytical platform: Agilent microarrays were normalized using the normalizeBetweenArrays function in limma (v3.44.3) and Affymetrix data were normalized using the rma function in affy (v1.66.0) packages. Microarray probe IDs were mapped to Gene symbol according to the GPL annotation files provided in NCBI. Probes mapped to multiple gene symbols were removed and genes mapped to multiple probe IDs were summarized by calculating the mean. The ComBat function in sva (v3.36.0) was then used to correct the technical batch effect. Differential expression analysis was performed using the limma package (version 3.40.6). Genes with adjusted P value (Benjamini–Hochberg) less than 0.05 were considered as differentially expressed genes (DEGs). RNA-seq data of control *vs*. IPF lung tissues were downloaded from GEO with the accession GSE92592 (32). Raw read counts were normalized and analyzed by using R package of DESeq2. Pathway enrichment analysis was performed in Metascape with default parameters. Adjusted *P* value < 0.05 as the cut-off criterion was considered statistically significant.

### Oxidative stress and HIF score calculation, and grouping

An oxidative stress signature was established previously based on upregulated genes in IPF cell populations from HALLMARK_REACTIVE_OXYGEN_SPECIES_PATHWAY gene set (*ABCC1, CDKN2D, FES, GCLC, GCLM, GLRX2, HHEX, IPCEF1, JUNB, LAMTOR5, LSP1, MBP, MGST1, MPO, NDUFA6, PFKP, PRDX1, PRDX2, PRDX4, PRNP, SBNO2, SCAF4, SOD1, SOD2, RXN1, TXN, TXNRD1*) (2). Buffa and colleagues summarized a HIF signature consisting of 15 genes (*ACOT7, ADM, ALDOA, CDKN3, ENO1, LDHA, MIF, MRPS17, NDRG1, P4HA1, PGAM1, SLC2A1, TPI1, TUBB6* and *VEGFA*) by combining gene function analysis and gene co-expression analysis in *vivo* (3). Based on these signatures, the oxidative stress score and HIF score for each sample were calculated by using gene set variation analysis in the GSVA (v1.36.2) package (7). To classify HIF scores, we employed unsupervised hierarchical clustering (ward.D) to cluster samples according to the HIF scores. Student’s *t*-test was used to evaluate the statistical difference between high *vs*. low score groups.

### Survival analysis

Univariate and multivariate Cox proportional hazard models were used to assess the hazard ratio (HR) of each parameter via the survminer (v0.4.8). We performed log-rank tests to compare Kaplan-Meier survival curves between groups with high or low scores by survival (v3.2-3). Prediction error curves of each prognostic model were generated from pec (v2019.11.03).

### Single-cell data analysis

The single-cell dataset was downloaded from GEO (Accession: GSE122960). Seurat (v2.2) was used to filter cells, cluster and perform t-distributed stochastic neighbor embedding (t-SNE) reduction and identify cell types by marker genes described by Reyfman and colleagues (10).

### CIBERSORTx analysis

We used CIBERSORTx to estimate the percentage of 22 types of infiltrating immune cells in each PBMC samples with default parameters (11). For BAL samples, the first step was to create the signature matrix of each cell type in IPF patients from single-cell gene expression profiling. The imputation of bulk BAL sample used the signature matrix with S-batch correction and 1000 permutations for significance analysis. Student’s *t*-test was used to evaluate the statistical difference in each cell population between the two conditions. *P*-values were adjusted for multiple testing using the Benjamini-Hochberg method.

### Statistics

Statistical analyses were performed in GraphPad Prism v7.02 (GraphPad Software Inc., San Diego, CA) unless otherwise indicated. We evaluated the correlations between oxidative stress score and HIF score using Pearson’s correlation. Normality of distribution was assessed using the D’Agostino-Pearson normality test. Statistical analyses of single comparisons of two groups utilised Student’s *t*-test or Mann-Whitney *U*-test for parametric and non-parametric data, respectively. Where appropriate, individual Student’s *t*-test results were corrected for multiple comparisons using the Holm-Sidak method. Results were considered significant if *P* < 0.05, where \**P* < 0.05, \*\**P* < 0.01, \*\*\**P* < 0.001, and \*\*\*\**P* < 0.0001.

## Data Availability

All data are provided within the manuscript. For bioinformatic analyses codes were implemented in R and have been deposited in GitHub: https://github.com/yz3n18/HIF_IPF.

## Code availability

Codes were implemented in R and have been deposited in GitHub: https://github.com/yz3n18/HIF_IPF.

## Acknowledgements

This project was supported by Medical Research Council [MR/S025480/1], an Academy of Medical Sciences/the Wellcome Trust Springboard Award [SBF002\1038] and the AAIR Charity. YZ was supported by an Institute for Life Sciences PhD Studentship.

## Supplementary Figures

**Figure 1-figure supplement 1.**
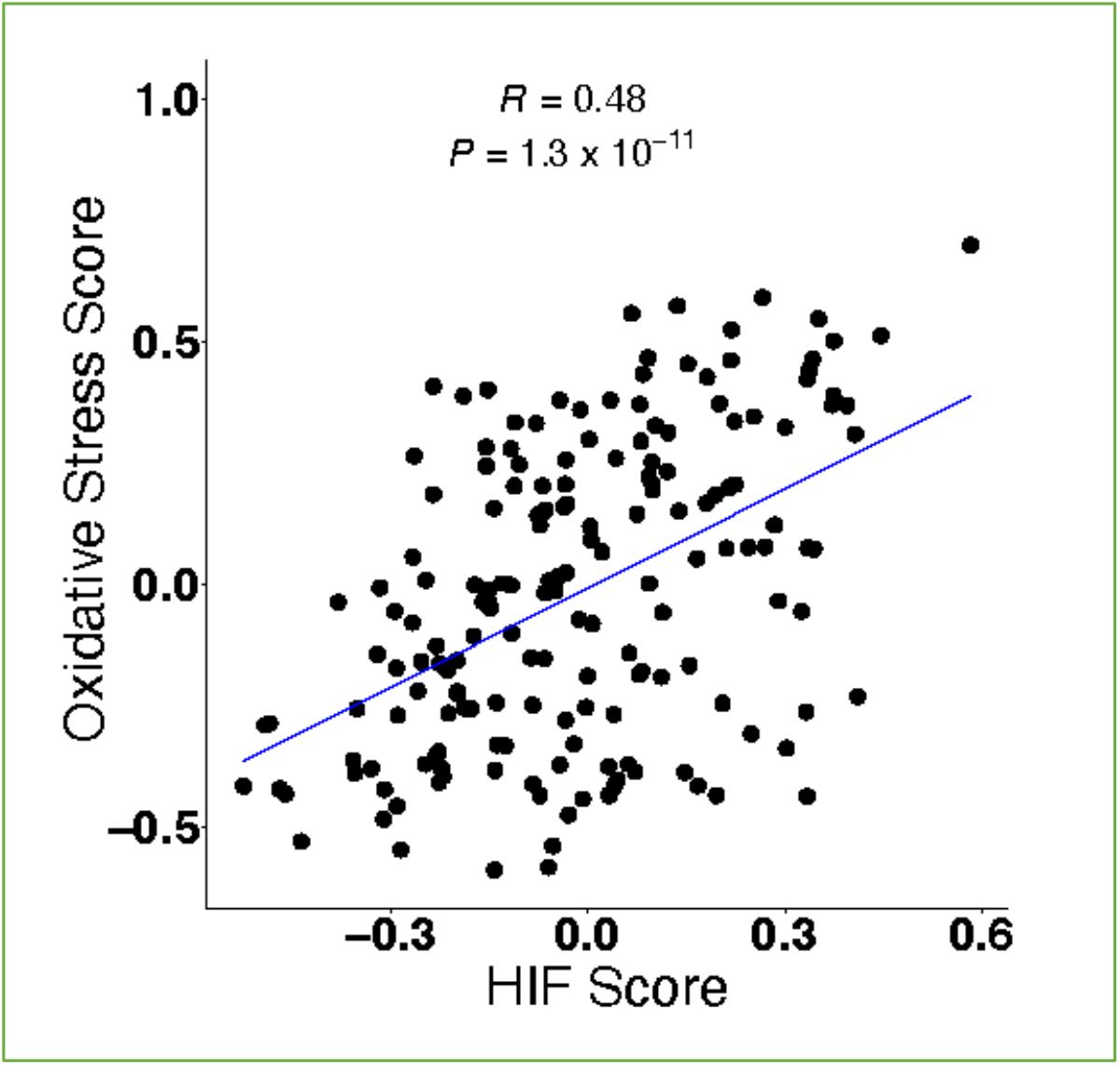
A validated HIF score is increased across tissue compartments in patients with IPF. The scatter plot for the correlation between HIF score and oxidative stress score calculated using GSVA in IPF BAL samples (GSE70867) (Pearson’s correlation *R* = 0.48 and *P* = 1.3×10^−11^).

**Figure 2-figure supplement 1.**
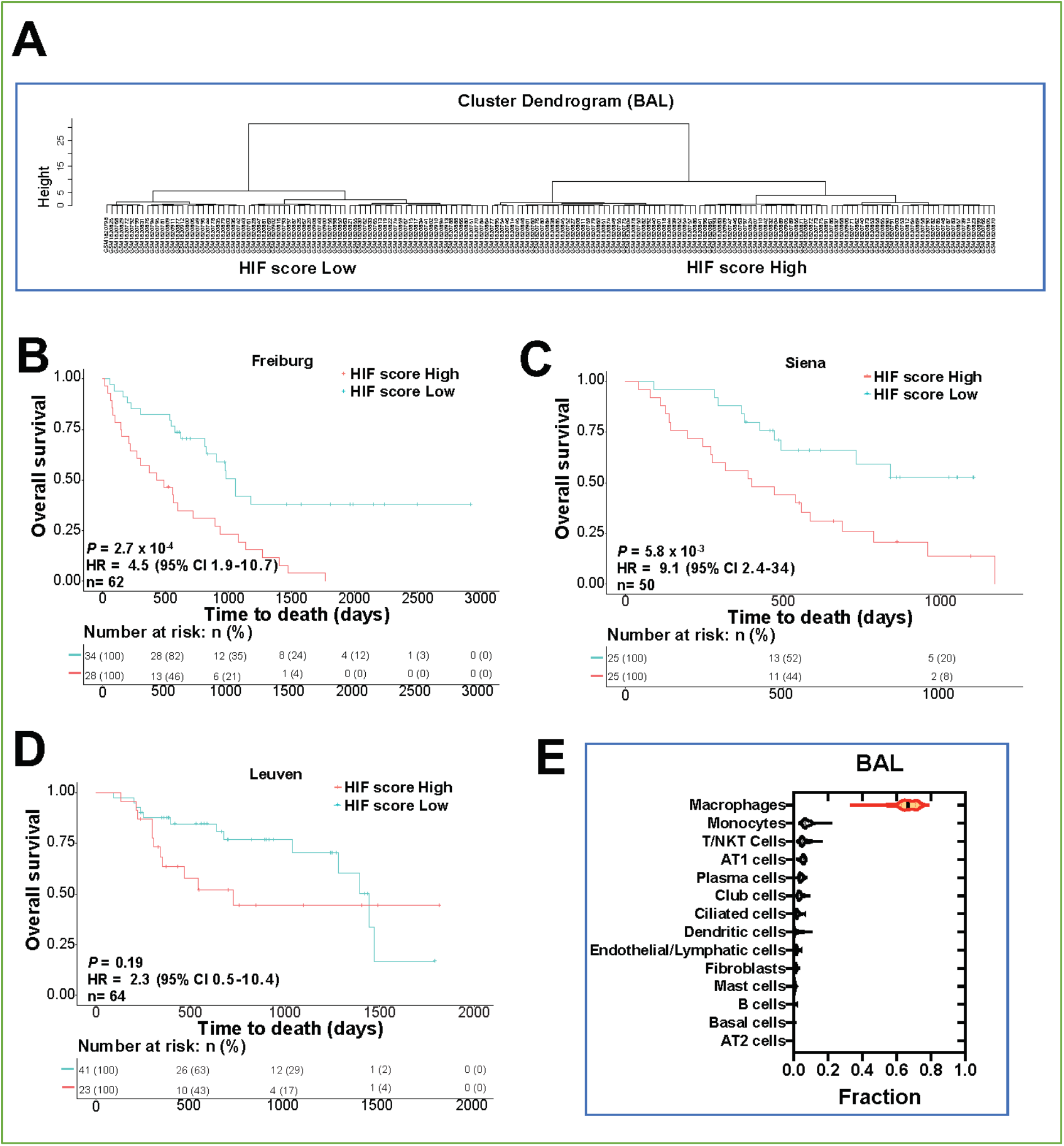
The HIF score in BAL predicts mortality in patients with IPF. (*A*) Hierarchical clustering dendrogram plot of BAL samples from IPF patients (GSE70867) according to HIF scores. (*B*-*D*) Kaplan-Meier plot showing the overall survival in IPF patients of each cohort with low *vs*. high HIF scores in BAL. Numbers below are *n* (%). *P* values, hazard ratio (HR), 95% confidence interval (CI) and patient number (*n*) are indicated.(*E*) Violin plot showing cell deconvolution proportions based on a signature matrix derived from the single-cell RNAseq found in Reyfman et al in BAL samples from IPF patients with (GSE70867)

**Figure 3-figure supplement 1.**
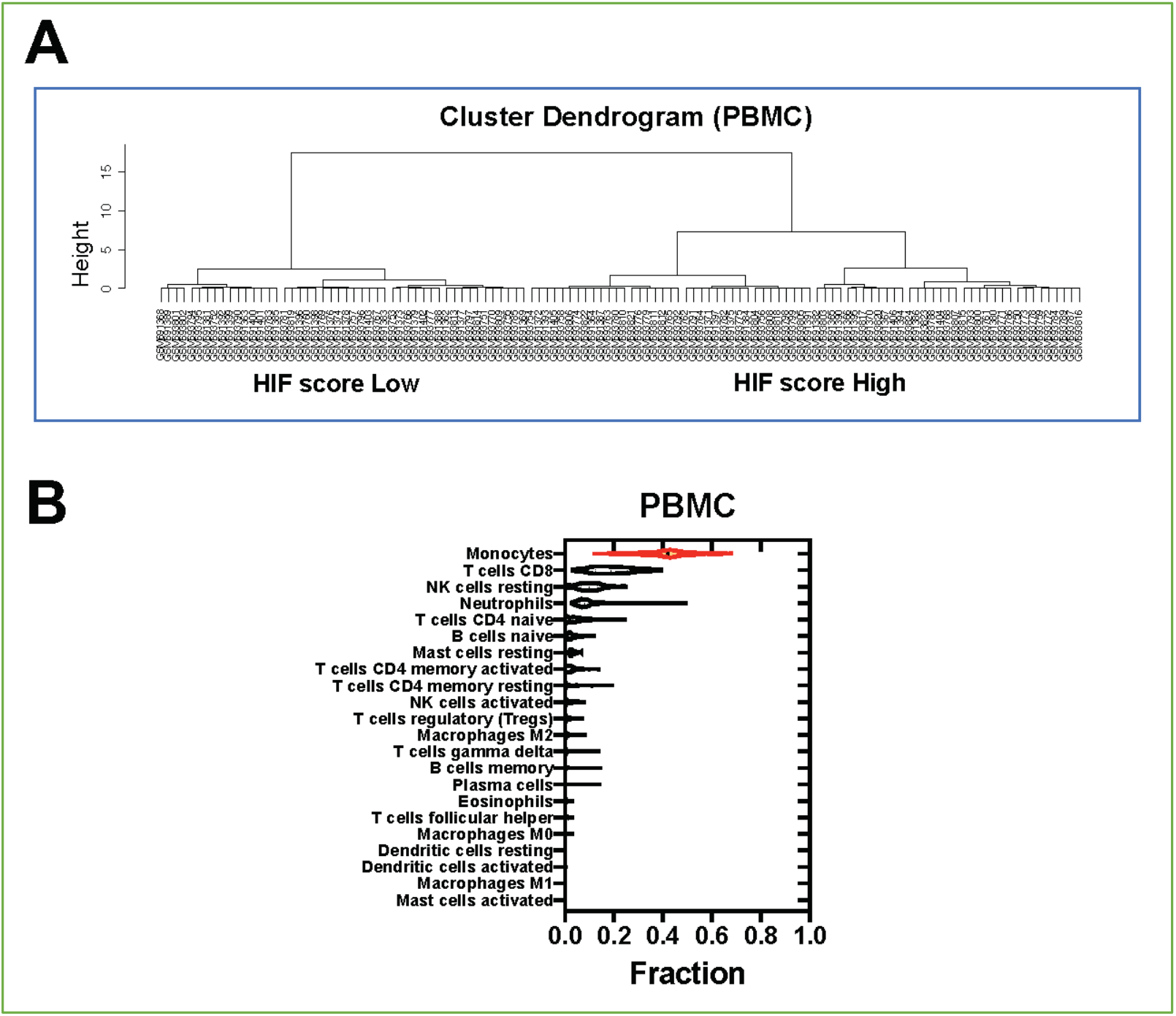
The HIF score is increased in PBMC from IPF patients. (*A*) Hierarchical clustering dendrogram plot of PBMC samples (GSE28221) from IPF patients according to HIF scores. (*B*) Violin plot showing cell compositions of PBMC using CIBERSORTx analysis.

**Figure 4-figure supplement 1.**
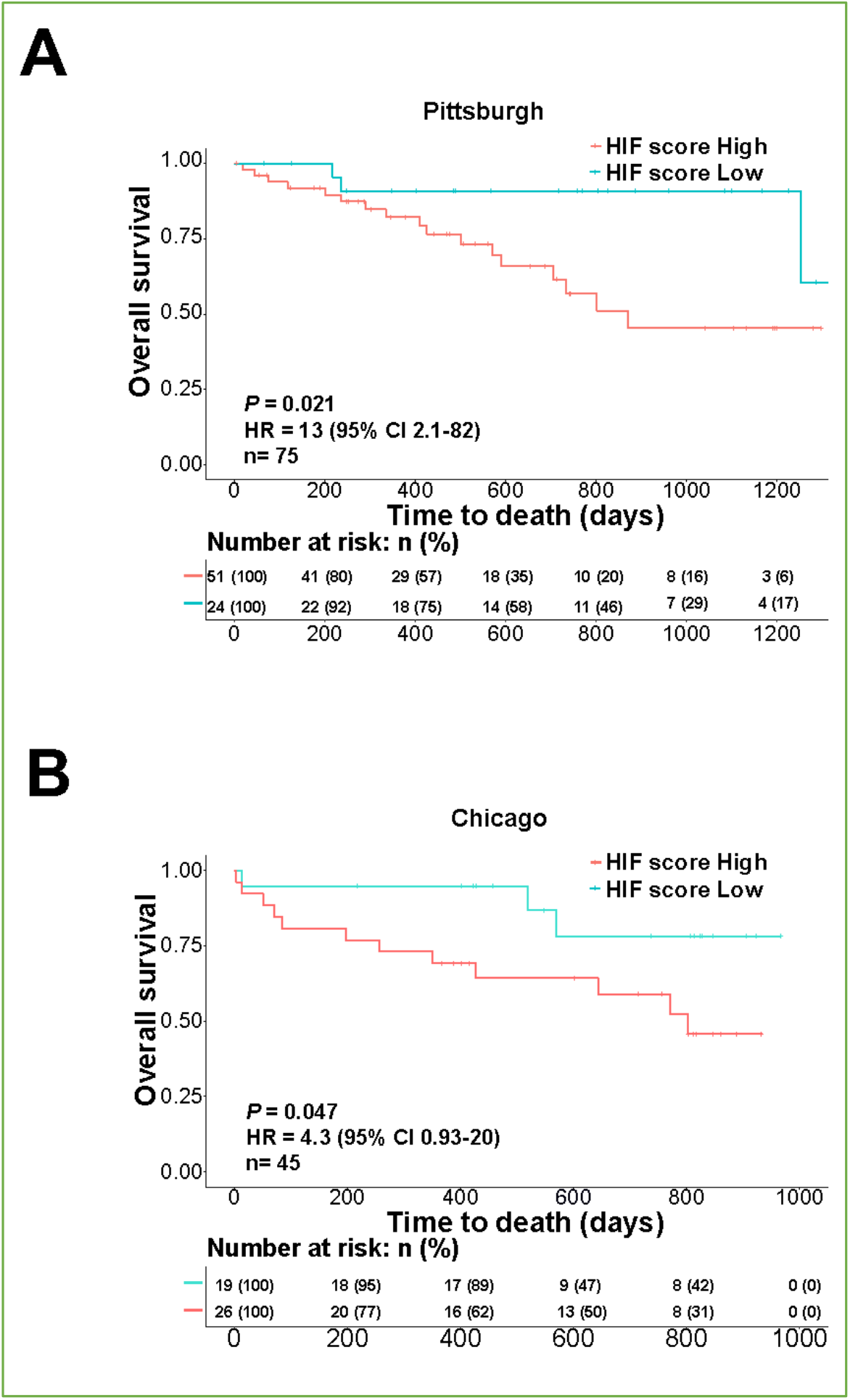
The HIF score in PBMC from IPF patients predicts mortality. Kaplan-Meier plot showing the overall survival in IPF patients of in (A) Pittsburgh and (B) Chicago cohorts with low *vs*. high HIF scores in PBMC (GSE28042 and GSE27957). Numbers below are *n* (%). *P* values, hazard ratio (HR), 95% confidence interval (CI) and patient number (*n*) are indicated.

## References

1. Jones MG, Andriotis OG, Roberts JJ, Lunn K, Tear VJ, Cao L, Ask K, Smart DE, Bonfanti A, Johnson P, Alzetani A, Conforti F, Doherty R, Lai CY, Johnson B, Bourdakos KN, Fletcher SV, Marshall BG, Jogai S, Brereton CJ, Chee SJ, Ottensmeier CH, Sime P, Gauldie J, Kolb M, Mahajan S, Fabre A, Bhaskar A, Jarolimek W, Richeldi L, O’Reilly KM, Monk PD, Thurner PJ, Davies DE. Nanoscale dysregulation of collagen structure-function disrupts mechano-homeostasis and mediates pulmonary fibrosis. Elife 2018; 7: e36354.

2. Brereton C, Yao L, Zhou Y, Vukmirovic M, Bell J, Ridley RA, Davies ER, Dean LSN, Andriotis OG, Conforti F, Mohammed S, Wallis T, Tavassoli A, Ewing R, Alzetani A, Marshall BG, Fletcher SV, Thurner PJ, Fabre A, Kaminski N, Richeldi L, Bhaskar A, Loxham M, Davies DE, Wang Y, Jones MG. Pseudohypoxic HIF pathway activation dysregulates collagen structure-function in human lung fibrosis. eLife 2022 Accepted/in press

3. Buffa FM, Harris AL, West CM, Miller CJ. Large meta-analysis of multiple cancers reveals a common, compact and highly prognostic hypoxia metagene. Br J Cancer 2010; 102: 428–435.

4. Ye Y, Hu Q, Chen H, Liang K, Yuan Y, Xiang Y, Ruan H, Zhang Z, Song A, Zhang H, Liu L, Diao L, Lou Y, Zhou B, Wang L, Zhou S, Gao J, Jonasch E, Lin SH, Xia Y, Lin C, Yang L, Mills GB, Liang H, Han L. Characterization of Hypoxia-associated Molecular Features to Aid Hypoxia-Targeted Therapy. Nat Metab 2019; 1: 431–444.

5. Bhandari V, Hoey C, Liu LY, Lalonde E, Ray J, Livingstone J, Lesurf R, Shiah YJ, Vujcic T, Huang X, Espiritu SMG, Heisler LE, Yousif F, Huang V, Yamaguchi TN, Yao CQ, Sabelnykova VY, Fraser M, Chua MLK, van der Kwast T, Liu SK, Boutros PC, Bristow RG. Molecular landmarks of tumor hypoxia across cancer types. Nat Genet 2019; 51: 308–318.

6. Bhandari V, Li CH, Bristow RG, Boutros PC, Consortium P. Divergent mutational processes distinguish hypoxic and normoxic tumours. Nat Commun 2020; 11: 737.

7. Hanzelmann S, Castelo R, Guinney J. GSVA: gene set variation analysis for microarray and RNA-seq data. BMC Bioinformatics 2013; 14: 7.

8. Prasse A, Binder H, Schupp JC, Kayser G, Bargagli E, Jaeger B, Hess M, Rittinghausen S, Vuga L, Lynn H, Violette S, Jung B, Quast K, Vanaudenaerde B, Xu Y, Hohlfeld JM, Krug N, Herazo-Maya JD, Rottoli P, Wuyts WA, Kaminski N. BAL Cell Gene Expression Is Indicative of Outcome and Airway Basal Cell Involvement in Idiopathic Pulmonary Fibrosis. American journal of respiratory and critical care medicine 2019; 199: 622–630.

9. Ley B, Ryerson CJ, Vittinghoff E, Ryu JH, Tomassetti S, Lee JS, Poletti V, Buccioli M, Elicker BM, Jones KD, King TE, Jr., Collard HR. A multidimensional index and staging system for idiopathic pulmonary fibrosis. Annals of internal medicine 2012; 156: 684–691.

10. Reyfman PA, Walter JM, Joshi N, Anekalla KR, McQuattie-Pimentel AC, Chiu S, Fernandez R, Akbarpour M, Chen CI, Ren Z, Verma R, Abdala-Valencia H, Nam K, Chi M, Han S, Gonzalez-Gonzalez FJ, Soberanes S, Watanabe S, Williams KJN, Flozak AS, Nicholson TT, Morgan VK, Winter DR, Hinchcliff M, Hrusch CL, Guzy RD, Bonham CA, Sperling AI, Bag R, Hamanaka RB, Mutlu GM, Yeldandi AV, Marshall SA, Shilatifard A, Amaral LAN, Perlman H, Sznajder JI, Argento AC, Gillespie CT, Dematte J, Jain M, Singer BD, Ridge KM, Lam AP, Bharat A, Bhorade SM, Gottardi CJ, Budinger GRS, Misharin AV. Single-Cell Transcriptomic Analysis of Human Lung Provides Insights into the Pathobiology of Pulmonary Fibrosis. Am J Respir Crit Care Med 2019; 199: 1517–1536.

11. Newman AM, Steen CB, Liu CL, Gentles AJ, Chaudhuri AA, Scherer F, Khodadoust MS, Esfahani MS, Luca BA, Steiner D, Diehn M, Alizadeh AA. Determining cell type abundance and expression from bulk tissues with digital cytometry. Nat Biotechnol 2019; 37: 773–782.

12. Maher TM. Blood-based Diagnosis of Idiopathic Pulmonary Fibrosis. Fantasy or Reality? American journal of respiratory and critical care medicine 2016; 194: 1182–1184.

13. Huang LS, Berdyshev EV, Tran JT, Xie L, Chen J, Ebenezer DL, Mathew B, Gorshkova I, Zhang W, Reddy SP, Harijith A, Wang G, Feghali-Bostwick C, Noth I, Ma SF, Zhou T, Ma W, Garcia JG, Natarajan V. Sphingosine-1-phosphate lyase is an endogenous suppressor of pulmonary fibrosis: role of S1P signalling and autophagy. Thorax 2015; 70: 1138–1148.

14. Huang LS, Mathew B, Li H, Zhao Y, Ma SF, Noth I, Reddy SP, Harijith A, Usatyuk PV, Berdyshev EV, Kaminski N, Zhou T, Zhang W, Zhang Y, Rehman J, Kotha SR, Gurney TO, Parinandi NL, Lussier YA, Garcia JG, Natarajan V. The mitochondrial cardiolipin remodeling enzyme lysocardiolipin acyltransferase is a novel target in pulmonary fibrosis. American journal of respiratory and critical care medicine 2014; 189: 1402–1415.

15. Herazo-Maya JD, Noth I, Duncan SR, Kim S, Ma SF, Tseng GC, Feingold E, Juan-Guardela BM, Richards TJ, Lussier Y, Huang Y, Vij R, Lindell KO, Xue J, Gibson KF, Shapiro SD, Garcia JG, Kaminski N. Peripheral blood mononuclear cell gene expression profiles predict poor outcome in idiopathic pulmonary fibrosis. Science translational medicine 2013; 5: 205ra136.

16. Huang Y, Ma SF, Vij R, Oldham JM, Herazo-Maya J, Broderick SM, Strek ME, White SR, Hogarth DK, Sandbo NK, Lussier YA, Gibson KF, Kaminski N, Garcia JG, Noth A functional genomic model for predicting prognosis in idiopathic pulmonary fibrosis. BMC pulmonary medicine 2015; 15: 147.

17. Newman AM, Liu CL, Green MR, Gentles AJ, Feng W, Xu Y, Hoang CD, Diehn M, Alizadeh AA. Robust enumeration of cell subsets from tissue expression profiles. Nature methods 2015; 12: 453–457.

18. Fox NS, Starmans MH, Haider S, Lambin P, Boutros PC. Ensemble analyses improve signatures of tumour hypoxia and reveal inter-platform differences. BMC Bioinformatics 2014; 15: 170.

19. Masson N, Singleton RS, Sekirnik R, Trudgian DC, Ambrose LJ, Miranda MX, Tian YM, Kessler BM, Schofield CJ, Ratcliffe PJ. The FIH hydroxylase is a cellular peroxide sensor that modulates HIF transcriptional activity. EMBO Rep 2012; 13: 251–257.

20. Idiopathic Pulmonary Fibrosis Clinical Research N, Martinez FJ, de Andrade JA, Anstrom KJ, King TE, Jr., Raghu G. Randomized trial of acetylcysteine in idiopathic pulmonary fibrosis. N Engl J Med 2014; 370: 2093–2101.

21. Feng F, Zhang J, Wang Z, Wu Q, Zhou X. Efficacy and safety of N-acetylcysteine therapy for idiopathic pulmonary fibrosis: An updated systematic review and meta-analysis. Exp Ther Med 2019; 18: 802–816.

22. Oldham JM, Ma SF, Martinez FJ, Anstrom KJ, Raghu G, Schwartz DA, Valenzi E, Witt L, Lee C, Vij R, Huang Y, Strek ME, Noth I, Investigators IP. TOLLIP, MUC5B, and the Response to N-Acetylcysteine among Individuals with Idiopathic Pulmonary Fibrosis. Am J Respir Crit Care Med 2015; 192: 1475–1482.

23. Scott MKD, Quinn K, Li Q, Carroll R, Warsinske H, Vallania F, Chen S, Carns MA, Aren K, Sun J, Koloms K, Lee J, Baral J, Kropski J, Zhao H, Herzog E, Martinez FJ, Moore BB, Hinchcliff M, Denny J, Kaminski N, Herazo-Maya JD, Shah NH, Khatri P. Increased monocyte count as a cellular biomarker for poor outcomes in fibrotic diseases: a retrospective, multicentre cohort study. The Lancet Respiratory medicine 2019; 7: 497–508.

24. Karampitsakos T, Torrisi S, Antoniou K, Manali E, Korbila I, Papaioannou O, Sampsonas F, Katsaras M, Vasarmidi E, Papakosta D, Domvri K, Fouka E, Organtzis I, Daniil Z, Dimeas I, Kirgou P, Gourgoulianis KI, Papanikolaou IC, Markopoulou K, Kounti G, Tsapakidou E, Papadopoulou E, Tatsis K, Gogali A, Kostikas K, Tzilas V, Chrysikos S, Papiris S, Bouros D, Kreuter M, Tzouvelekis A. Increased monocyte count and red cell distribution width as prognostic biomarkers in patients with Idiopathic Pulmonary Fibrosis. Respir Res 2021; 22: 140.

25. Kreuter M, Lee JS, Tzouvelekis A, Oldham JM, Molyneaux PL, Weycker D, Atwood M, Kirchgaessler KU, Maher TM. Monocyte Count as a Prognostic Biomarker in Patients with Idiopathic Pulmonary Fibrosis. Am J Respir Crit Care Med 2021; 204: 74–81.

26. Semba H, Takeda N, Isagawa T, Sugiura Y, Honda K, Wake M, Miyazawa H, Yamaguchi Y, Miura M, Jenkins DM, Choi H, Kim JW, Asagiri M, Cowburn AS, Abe H, Soma K, Koyama K, Katoh M, Sayama K, Goda N, Johnson RS, Manabe I, Nagai R, Komuro I. HIF-1alpha-PDK1 axis-induced active glycolysis plays an essential role in macrophage migratory capacity. Nat Commun 2016; 7: 11635.

27. Alvarez-Martins I, Remedio L, Matias I, Diogo LN, Monteiro EC, Dias S. The impact of chronic intermittent hypoxia on hematopoiesis and the bone marrow microenvironment. Pflugers Archiv : European journal of physiology 2016; 468: 919–932.

28. Rathinam C, Poueymirou WT, Rojas J, Murphy AJ, Valenzuela DM, Yancopoulos GD, Rongvaux A, Eynon EE, Manz MG, Flavell RA. Efficient differentiation and function of human macrophages in humanized CSF-1 mice. Blood 2011; 118: 3119–3128.

29. Roiniotis J, Dinh H, Masendycz P, Turner A, Elsegood CL, Scholz GM, Hamilton JA. Hypoxia prolongs monocyte/macrophage survival and enhanced glycolysis is associated with their maturation under aerobic conditions. Journal of immunology 2009; 182: 7974–7981.

30. Fraser E, Denney L, Antanaviciute A, Blirando K, Vuppusetty C, Zheng Y, Repapi E, Iotchkova V, Taylor S, Ashley N, St Noble V, Benamore R, Hoyles R, Clelland C, Rastrick JMD, Hardman CS, Alham NK, Rigby RE, Simmons A, Rehwinkel J, Ho LP. Multi-Modal Characterization of Monocytes in Idiopathic Pulmonary Fibrosis Reveals a Primed Type I Interferon Immune Phenotype. Front Immunol 2021; 12: 623430.

31. Chaturvedi P, Gilkes DM, Takano N, Semenza GL. Hypoxia-inducible factor-dependent signaling between triple-negative breast cancer cells and mesenchymal stem cells promotes macrophage recruitment. Proc Natl Acad Sci U S A 2014; 111: E2120–2129.

32. Schafer MJ, White TA, Iijima K, Haak AJ, Ligresti G, Atkinson EJ, Oberg AL, Birch J, Salmonowicz H, Zhu Y, Mazula DL, Brooks RW, Fuhrmann-Stroissnigg H, Pirtskhalava T, Prakash YS, Tchkonia T, Robbins PD, Aubry MC, Passos JF, Kirkland JL, Tschumperlin DJ, Kita H, LeBrasseur NK. Cellular senescence mediates fibrotic pulmonary disease. Nat Commun 2017; 8: 14532.

